# Predictors of hospitalisation and death due to SARS-CoV-2 infection in Finland: a population-based register study with implications to vaccinations

**DOI:** 10.1101/2021.07.04.21259954

**Authors:** Heini Salo, Toni Lehtonen, Kari Auranen, Ulrike Baum, Tuija Leino

**Affiliations:** Infectious Disease Control and Vaccinations Unit, Department of Health Security, Finnish Institute for Health and Welfare (THL), Helsinki, Finland; The Center of Statistics, University of Turku, Turku, Finland

**Keywords:** SARS-CoV-2, COVID-19, risk factors, elderly, chronically ill

## Abstract

**Introduction:** The aim of this study was to investigate how age and underlying medical conditions affect the risk of severe outcomes following SARS-CoV-2 infection and how they should be weighed while prioritising vaccinations against COVID-19.

**Methods:** This population-based register study includes all SARS-CoV-2 PCR-test-positive cases until 24 Feb 2021, based on the Finnish National Infectious Diseases Register. The cases were linked to other registers to identify presence of comorbidities and severe outcomes (hospitalisation, intensive care treatment, death). The odds of severe outcomes were compared in those with and without the pre-specified comorbidities using logistic regression. Furthermore, population-based rates were compared between those with a given comorbidity and those without any of the specified comorbidities using negative binomial regression.

**Results:** Age and various comorbidities were found to be predictors of severe COVID-19. Compared to 60–69-year-olds, the odds ratio (OR) of death was 7.1 for 70–79-year-olds, 26.7 for 80–89-year-olds, and 55.8 for ≥90-year-olds. Among the 20–69-year-olds, chronic renal disease (OR 9.4), malignant neoplasms (5.8), hematologic malignancy (5.6), chronic pulmonary disease (5.4), and cerebral palsy or other paralytic syndromes (4.6) were strongly associated with COVID-19 mortality; severe disorders of the immune system (8.0), organ or stem cell transplant (7.2), chronic renal disease (6.7), and diseases of myoneural junction and muscle (5.5) were strongly associated with COVID-19 hospitalisation. Type 2 diabetes and asthma, two very common comorbidities, were associated with all three outcomes, with ORs from 2.1 to 4.3. The population-based rate of SARS-CoV-2 infection decreased with age. Taking the 60–69-year-olds as reference, the rate ratio was highest (3.0) for 20–29-year-olds but <1 for 70–79-year-olds and 80–89-year-olds.

**Conclusion:** Comorbidities predispose for severe COVID-19 among younger ages. In vaccine prioritisation both the risk of infection and the risk of severe outcomes, if infected, should be combined.

## Introduction

Already during summer 2020, the first listings of predisposing conditions for severe COVID-19 were published to inform the public on the need to shield the most vulnerable.[1–6] Furthermore, while efficacy studies of COVID-19 vaccines were still being carried out, more detailed research was published on predisposing factors for severe infection outcomes leading to hospitalisation [7,8], intensive care [7,9,10], or death [7,8,11–14]. The implications from these studies have been valuable in planning how to allocate the limited number of vaccine doses optimally to prevent hospitalisations, need to intensive care and deaths.[15] For example, one of the questions was whether the elderly or individuals with certain background illnesses should be prioritised for COVID-19 vaccination.[15] Along with the other European Union countries, Finland joined a common procurement system and subsequently commenced COVID-19 vaccinations in late December 2020.

We carried out a country-level analysis of factors predisposing for severe COVID-19 outcomes, based on the possibility to link information across several national health care registers using the unique personal identifier. The specific aim of our study was to investigate how age and presence of underlying medical conditions affect the risk of severe COVID-19 outcomes and how age and medical conditions should be weighed against each other while prioritising vaccination. We thus concentrated on assessing the risk of hospitalisation, intensive care treatment and death in those with SARS-CoV-2 infection, with specific attention to the elderly and persons with certain chronical illnesses and those taking potentially predisposing medications. We here report the results with a particular focus on how these data were used to support effective COVID-19 vaccination intervention in Finland.

## Material and Methods

### Data sources and case definitions

The total population of Finland in 2019 was 5.5 million, 51% of which were women. Age- and gender-specific population sizes were obtained from the Finnish Population Information System, a computerised national database which registers individual-based information for all Finnish residents, including name, gender, personal identity code, address, date of birth, and death. The personal identity code remains unchanged throughout the lifetime and is used for patient identification in all health care registers in Finland.

We established a dataset comprising virtually the entire Finnish population by linking data from multiple population-based and nationwide registers. This COVID-19 dataset included all individuals with a positive PCR test for SARS-CoV-2 identified between 1 January 2020 and 24 February 2021, based on the Finnish National Infectious Diseases Register. The SARS-CoV-2 cases were linked to other registers in order to identify severe outcomes. Three severe COVID-19 outcomes were defined as inpatient hospitalisation (Care Register for Health Care), admission to intensive care unit (ICU; the Finnish Intensive Care Consortium’s Database, FICC), and death within 30 days after the positive PCR test. All cases hospitalised 14 days before or after the positive PCR test with a diagnosis indicating respiratory infection, were considered as COVID-19 outcomes.

Individuals with chronic diseases or medical conditions (from now on comorbidities) known as general risk factors of infectious diseases or with risk factors for severe COVID-19 from published studies were identified from the registers using a pre-defined list of diagnoses and codes (Table 1). The following registers were employed: the Care Register for Health Care, the Register of Primary Health Care Visits, as well as the Special Reimbursement Register for Medicine Expenses and Prescription Centre database, both maintained by the Social Insurance Institution of Finland (SII). The dataset covers for the most part all health care providers in Finland. The dataset was linked to the Finnish Population Information System in order to identify individuals without any of the comorbidities potentially predisposing to severe COVID-19 listed in Table 1.

**Table 1.**
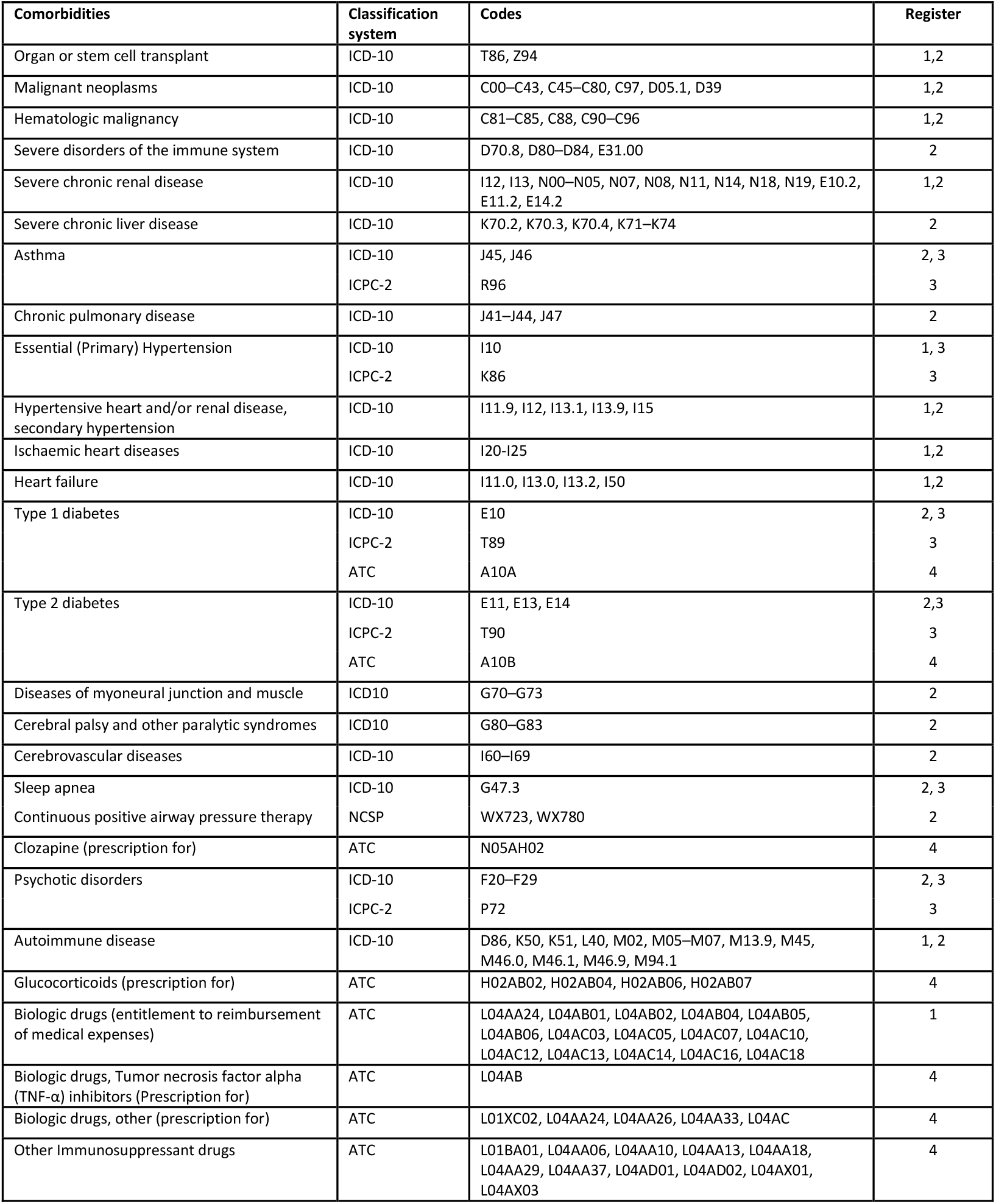
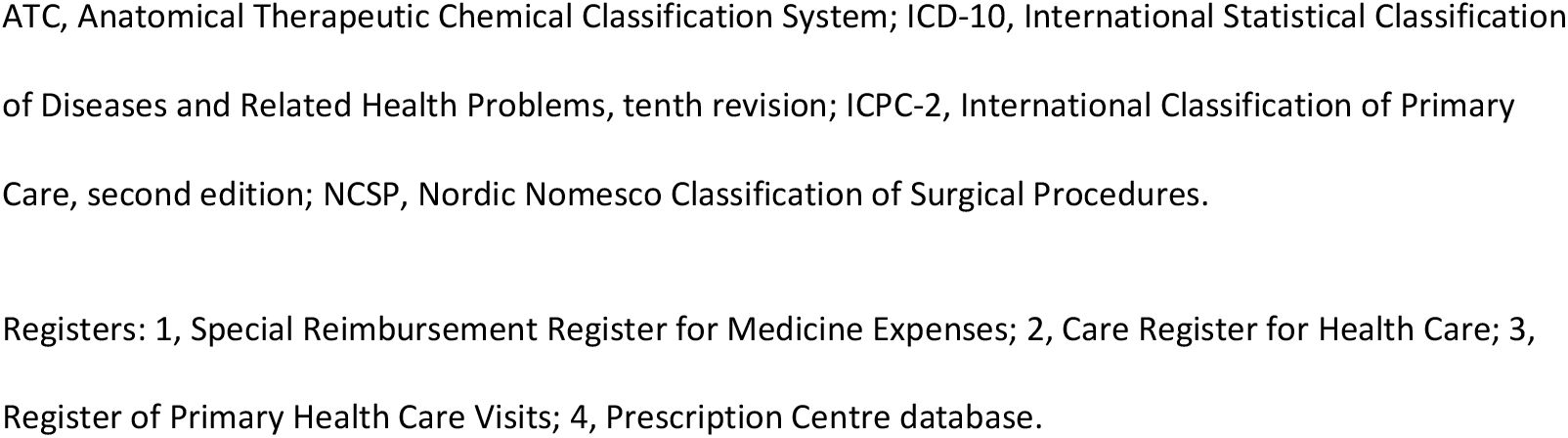
Comorbidities identified as potential risk factors predisposing to severe COVID-19

| Comorbidities | Classification system | Codes | Register |
| --- | --- | --- | --- |
| Organ or stem cell transplant | ICD-10 | T86, Z94 | 1,2 |
| Malignant neoplasms | ICD-10 | C00–C43, C45–C80, C97, D05.1, D39 | 1,2 |
| Hematologic malignancy | ICD-10 | C81–C85, C88, C90–C96 | 1,2 |
| Severe disorders of the immune system | ICD-10 | D70.8, D80–D84, E31.00 | 2 |
| Severe chronic renal disease | ICD-10 | I12, I13, N00–N05, N07, N08, N11, N14, N18, N19, E10.2, E11.2, E14.2 | 1,2 |
| Severe chronic liver disease | ICD-10 | K70.2, K70.3, K70.4, K71–K74 | 2 |
| Asthma | ICD-10 | J45, J46 | 2, 3 |
|  | ICPC-2 | R96 | 3 |
| Chronic pulmonary disease | ICD-10 | J41–J44, J47 | 2 |
| Essential (Primary) Hypertension | ICD-10 | I10 | 1, 3 |
|  | ICPC-2 | K86 | 3 |
| Hypertensive heart and/or renal disease, secondary hypertension | ICD-10 | I11.9, I12, I13.1, I13.9, I15 | 1,2 |
| Ischaemic heart diseases | ICD-10 | I20–I25 | 1,2 |
| Heart failure | ICD-10 | I11.0, I13.0, I13.2, I50 | 1,2 |
| Type 1 diabetes | ICD-10 | E10 | 2, 3 |
|  | ICPC-2 | T89 | 3 |
|  | ATC | A10A | 4 |
| Type 2 diabetes | ICD-10 | E11, E13, E14 | 2,3 |
|  | ICPC-2 | T90 | 3 |
|  | ATC | A10B | 4 |
| Diseases of myoneural junction and muscle | ICD10 | G70–G73 | 2 |
| Cerebral palsy and other paralytic syndromes | ICD10 | G80–G83 | 2 |
| Cerebrovascular diseases | ICD-10 | I60–I69 | 2 |
| Sleep apnea | ICD-10 | G47.3 | 2, 3 |
| Continuous positive airway pressure therapy | NCSP | WX723, WX780 | 2 |
| Clozapine (prescription for) | ATC | N05AH02 | 4 |
| Psychotic disorders | ICD-10 | F20–F29 | 2, 3 |
|  | ICPC-2 | P72 | 3 |
| Autoimmune disease | ICD-10 | D86, K50, K51, L40, M02, M05–M07, M13.9, M45, M46.0, M46.1, M46.9, M94.1 | 1, 2 |
| Glucocorticoids (prescription for) | ATC | H02AB02, H02AB04, H02AB06, H02AB07 | 4 |
| Biologic drugs (entitlement to reimbursement of medical expenses) | ATC | L04AA24, L04AB01, L04AB02, L04AB04, L04AB05, L04AB06, L04AC03, L04AC05, L04AC07, L04AC10, L04AC12, L04AC13, L04AC14, L04AC16, L04AC18 | 1 |
| Biologic drugs, Tumor necrosis factor alpha (TNF-α) inhibitors (Prescription for) | ATC | L04AB | 4 |
| Biologic drugs, other (prescription for) | ATC | L01XC02, L04AA24, L04AA26, L04AA33, L04AC | 4 |
| Other Immunosuppressant drugs | ATC | L01BA01, L04AA06, L04AA10, L04AA13, L04AA18, L04AA29, L04AA37, L04AD01, L04AD02, L04AX01, L04AX03 | 4 |

ATC, Anatomical Therapeutic Chemical Classification System; ICD-10, International Statistical Classification of Diseases and Related Health Problems, tenth revision; ICPC-2, International Classification of Primary Care, second edition; NCSP, Nordic Nomesco Classification of Surgical Procedures.
Registers: 1, Special Reimbursement Register for Medicine Expenses; 2, Care Register for Health Care; 3, Register of Primary Health Care Visits; 4, Prescription Centre database.

Individuals entitled to reimbursement of medicine expenses from 1 January 2020 onwards were identified from the SII register for Special Reimbursements for Medicine Expenses. In Finland, all residents entitled to special reimbursement meet a set of specified medical criteria.

Individuals using medication predisposing to immunodeficiency or using medication for type 1 or 2 diabetes were identified from the Prescription Centre database maintained by SII. Persons that had any of the prescribed medications of interest not more than two years prior to testing positive for SARS-CoV-2 were identified based on the Anatomical Therapeutic Chemical (ATC) codes.

We identified individuals who had used health care services due to any of the specified comorbidities not more than two years (cancer) or five years (all other comorbidities) prior to the positive PCR test. Individuals who had used secondary health care services due the specified comorbidities from the Care Register for Health Care with the International Classification of Diseases 10th revision (ICD-10) codes and the Nordic Classification of Surgical Procedures (NCSP) codes. Individuals who had used primary health care services were identified from the Register of Primary Health Care Visits with ICD-10 codes and the International Classification of Primary Care Second edition (ICPC-2) codes.

### Statistical analysis

We estimate the incidence of SARS-CoV-2 infection and severe COVID-19 outcomes by age, sex and comorbidity in individuals with a SARS-CoV-2 infection and in the Finnish population as a whole. The three severe outcomes were hospitalisation, admission to ICU, and death. We first estimated age-specific odds ratios (ORs) for each outcome (e.g. hospitalisation) in SARS-CoV-2 cases using logistic regression and adjusting for sex. We next estimated odds ratios, comparing the odds of the outcome (e.g. hospitalisation) in SARS-CoV-2 cases with and without each of the comorbidities, adjusting for age and sex. In addition, negative binomial regression was used to estimate rate ratios (RR) for SARS-CoV-2 infection and the three severe outcomes, comparing individuals with each of the comorbidities with individuals without any of the listed comorbidities (Table 1). The negative binomial model was used instead of Poisson regression to account for overdispersion in the rates. The analysis was adjusted for age and sex.

## Results

### Characteristics of the study population

In Finland, a total of 52 502 SARS-CoV-2 cases occurred from 1 March 2020 through 24 February 2021, 2743 (5.2%) of which were hospitalised, 538 (1.0%) were admitted to ICU, and 801 (1.5%) died within 30 days after the positive PCR-test (Table 2). The corresponding incidence rates per 100 000 person-years were 822.4 for SARS-CoV-2 cases, 43 for hospitalised cases, 8.4 for ICU cases and 12.5 for fatal cases. When restricting to 20–69-year-olds, there were 38 728 SARS-CoV-2 cases, 1 916 (4.9%) of which were hospitalised, 403 (1.0%) were admitted to ICU and 98 (0.3%) died within 30 days after the positive PCR-test.

**Table 2.**
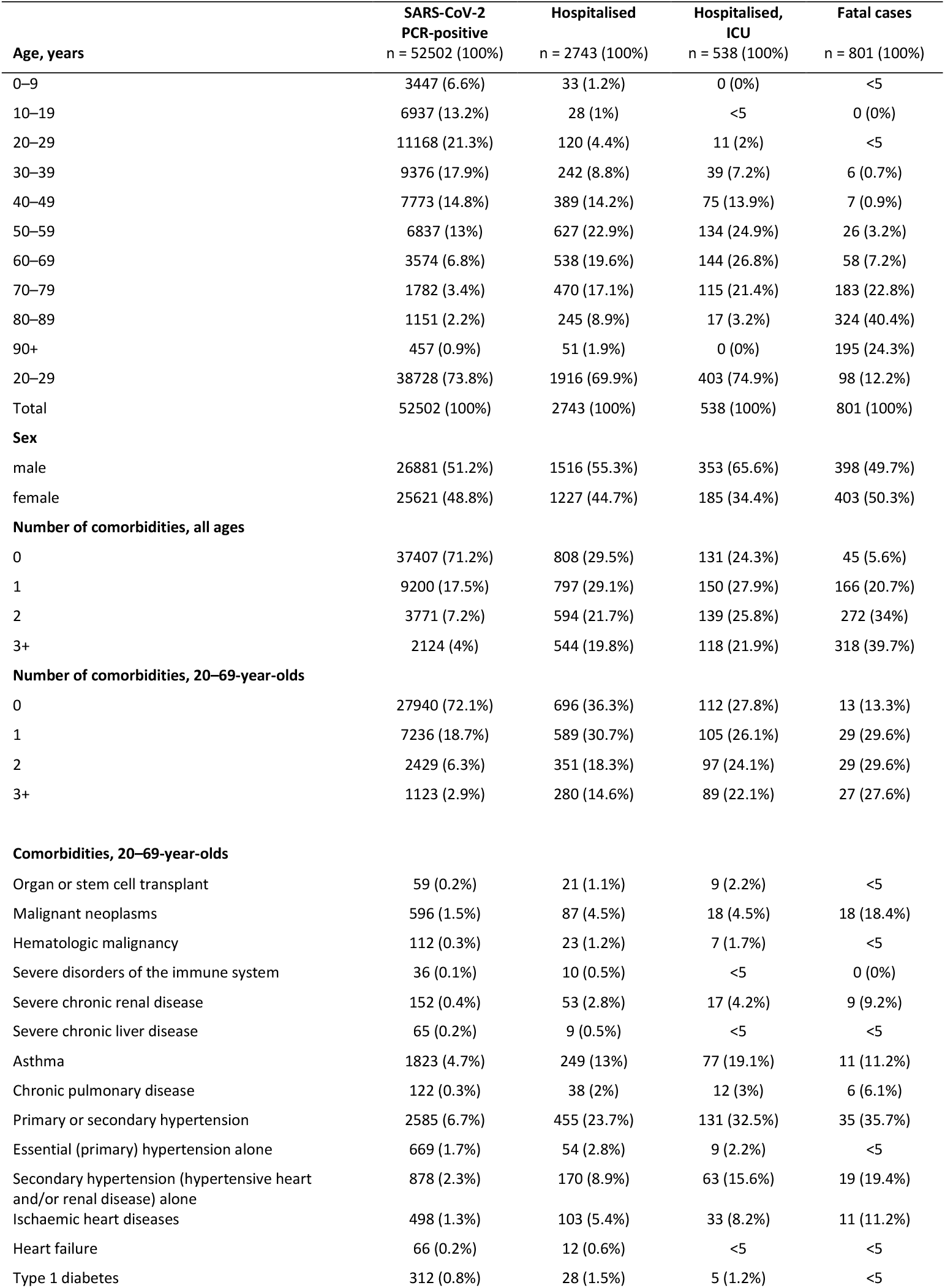
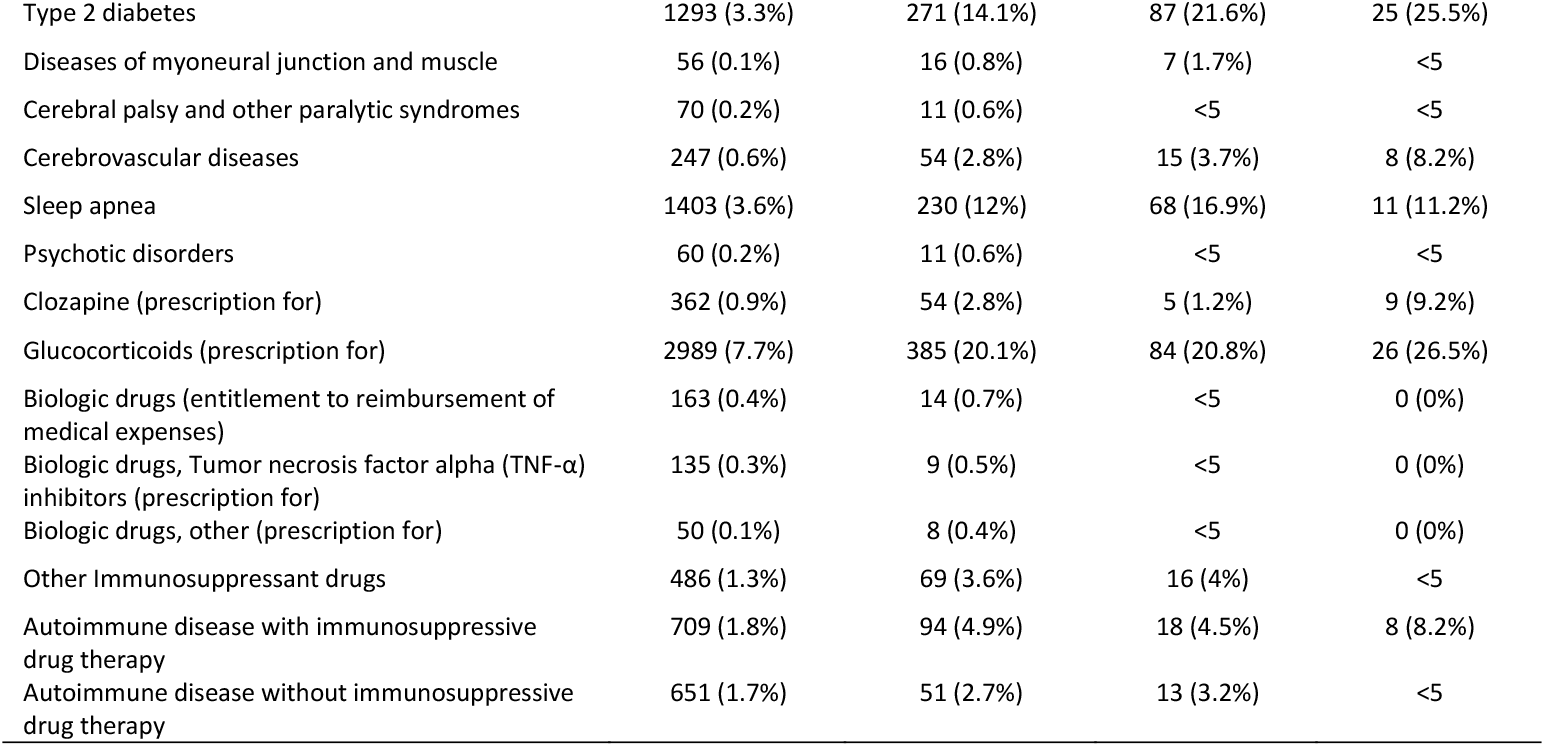
Baseline characteristics of cases of SARS-CoV-2 infection

The proportion of fatalities among SARS-CoV-2 cases increased with age (Figure 1). In those aged 60–69 years, all fatal cases but one had at least one predisposing factor for severe COVID-19. By contrast, in those aged ≥70 fatalities occurred also in individuals without any predisposing factors. In 20–69-year-olds, 28% of SARS-CoV-2 cases, 64% of hospitalised cases, 72% of ICU cases, and 87% of fatal cases had at least one predisposing factor.

**Figure 1.**
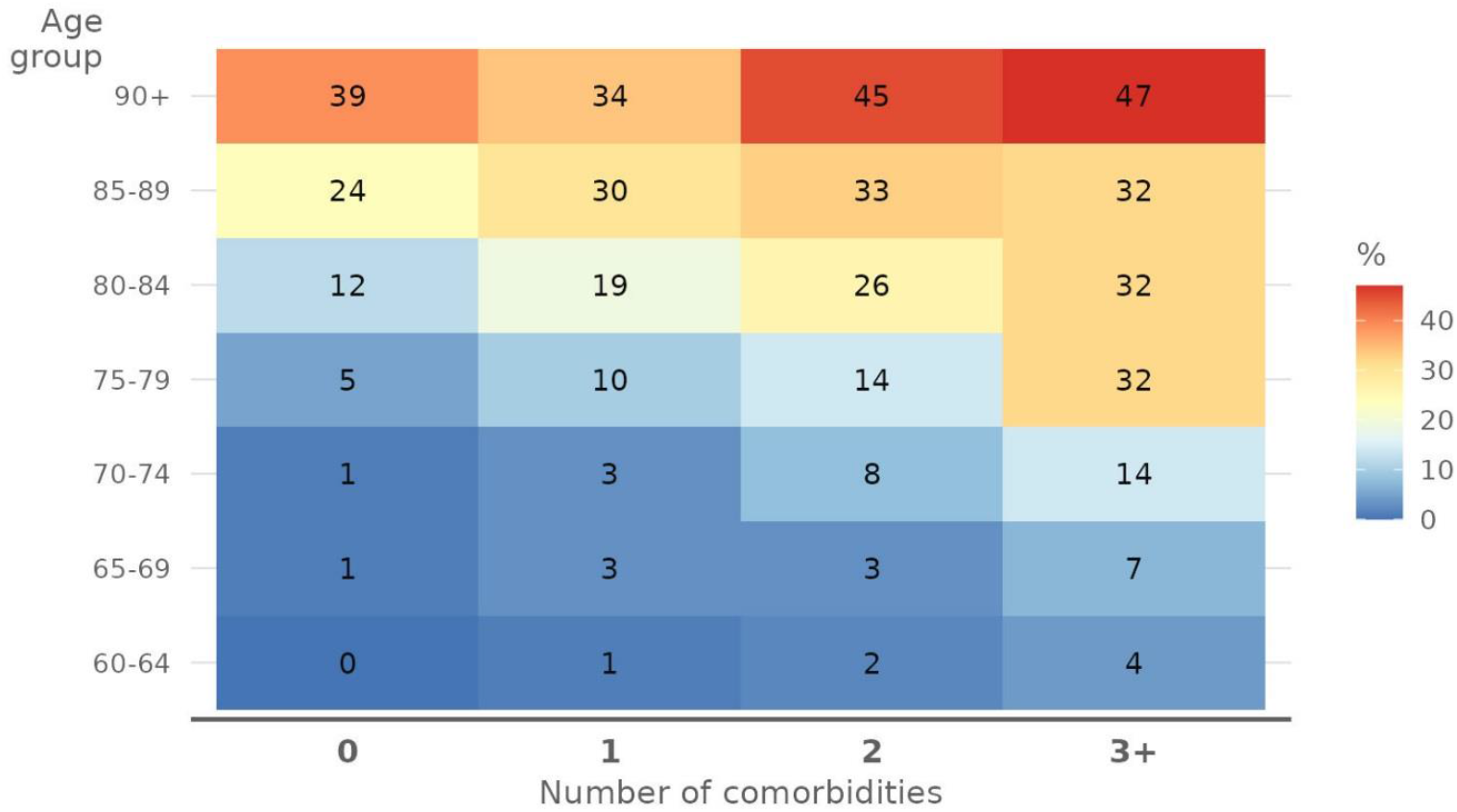
Proportion (%) of fatal outcome in cases of SARS-CoV-2 infection by age group and number of comorbidities.

The proportion of fatalities among SARS-CoV-2 cases increased with the number of comorbidities (Figure 1). In all age groups, the more comorbidities there were, the higher the mortality was among the infected. With three or more comorbidities the proportion of fatal cases varied from 4% (65–69 years) to 32% (75–89 years). In the oldest the proportion of fatal cases was even higher, irrespective of the number of comorbidities.

During the study period, SARS-CoV-2 infections were most frequent among 20–29-year-olds, hospitalised cases among 50–59-year-olds, ICU cases among 60–69-year-olds, and fatal cases among 80–89-year-olds (Table 2). The age-specific incidence rates per 100 000 person-years peaked for SARS-CoV-2 infections in those aged 20–29 (1475), for hospitalised cases in those aged 80–89 (79.6), for ICU cases in those aged 60–69 (17.6) and for fatal cases in those aged ≥90 (289.1) (Figure 2). Severe COVID-19 outcomes were thus clearly over-represented among the elderly. Only 6% (3390/52502) of all SARS-CoV-2 cases were detected in individuals aged ≥70 (Table 2). Yet 28% (766/2 743) of hospitalised cases, 25% (132/538) of ICU cases and 88% (702/801) of the fatal cases occurred in this age group.

**Figure 2.**
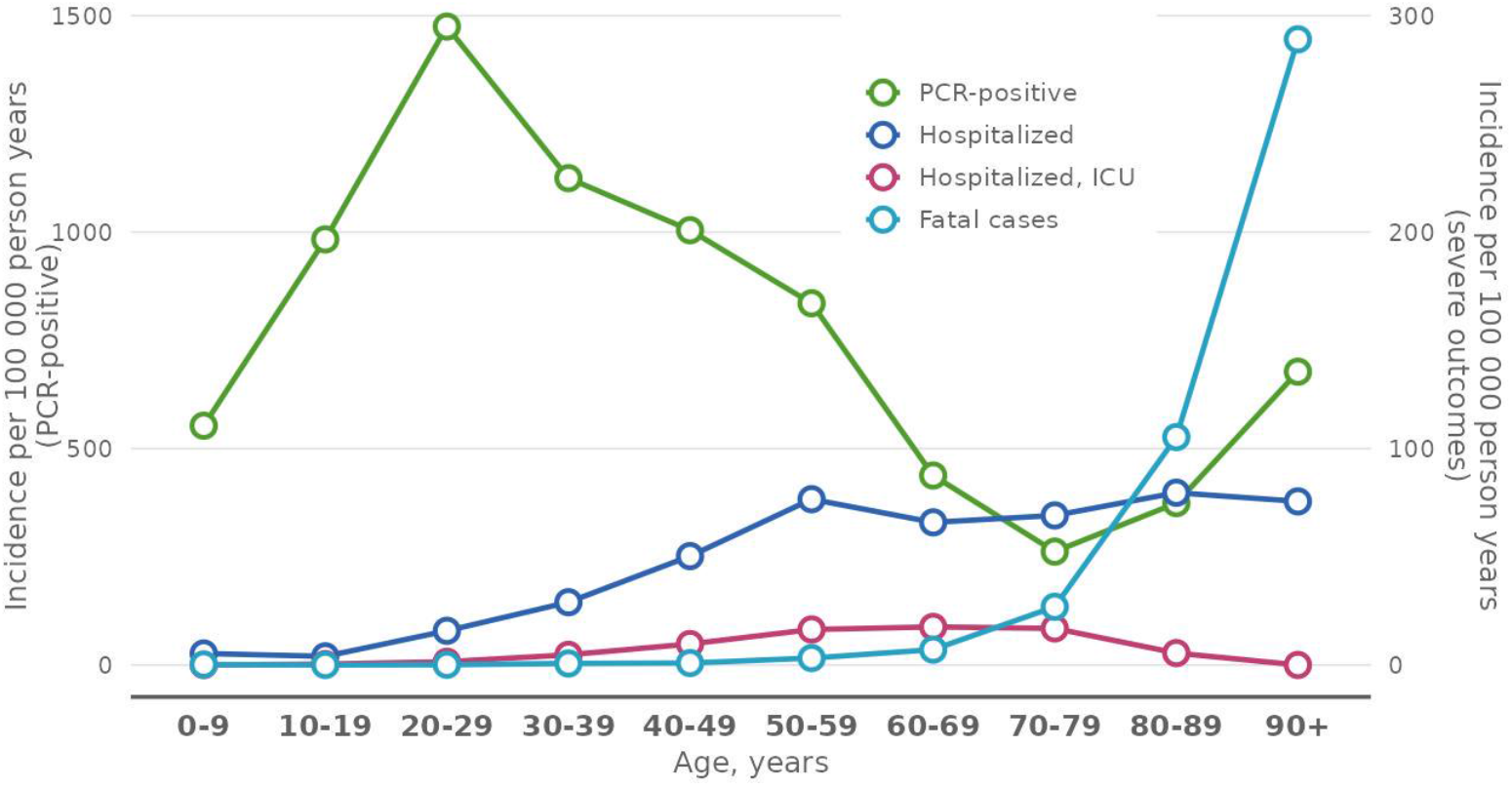
Incidence (per 100 000 person-years) of the SARS-Cov-2 infection and COVID-19 outcomes by age group in the Finnish population. SARS-Cov-2 cases on the left side Y axis and the remainder on right side Y axis.

### Predictors of hospitalisation, ICU admission and death

According to the logistic regression analysis, the risk of severe COVID-19 outcomes in an individual infected with SARS-CoV-2 increased with age. Compared to 60–69-year-olds, the odds ratio (OR) of death, adjusted for sex, was 7.1 (95% CI 5.3, 9.7) for the 70–79-year-olds, 26.7 (95% CI 20, 36) for the 80–89-year-olds, and 55.8 (95% CI 40.5, 77.9) for the ≥90-year-olds (Table 3a). Furthermore, the OR of hospitalisation was 2.0 (95% 1.8, 2.3) and of ICU admission 1.7 (95% CI 1.3, 2.2) for those aged 70–79 (Table 3a). In addition, male sex was identified as a risk factor for all severe outcomes: hospitalisation (OR 1.3; 95% CI 1.2, 1.4), ICU admission (OR 1.9; 95% CI 1.6, 2.3), and death (OR 1.9; 95% CI 1.6, 2.2).

**Table 3a.** Relative risk of severe COVID-19 (hospitalisation, ICU and death) in cases of SARS-Cov-2 infection by age group and sex. The reference category is 60–69-year-old females. Relative risks are presented in terms of odds ratios (OR) and their 95% confidence intervals (CI 95%).

|  | OR (95% CI) |  |  |  |  |  |
| --- | --- | --- | --- | --- | --- | --- |
|  | Hospitalised |  | Hospitalised, ICU |  | Fatal cases |  |
|  | Crude | Adjusted for sex | Crude | Adjusted for sex | Crude | Adjusted for sex |
| Age, years |  |  |  |  |  |  |
| 0–9 | 0.1 (0, 0.1) | 0.1 (0, 0.1) | 0 (0, 0) | 0 (0, 0) | 0 (0, 0.1) | 0 (0, 0.1) |
| 10–19 | 0 (0, 0) | 0 (0, 0) | 0 (0, 0) | 0 (0, 0) | 0 (0, 0) | 0 (0, 0) |
| 20–29 | 0.1 (0, 0.1) | 0.1 (0, 0.1) | 0 (0, 0) | 0 (0, 0) | 0 (0, 0) | 0 (0, 0) |
| 30–39 | 0.1 (0.1, 0.2) | 0.1 (0.1, 0.2) | 0.1 (0.1, 0.1) | 0.1 (0.1, 0.1) | 0 (0, 0.1) | 0 (0, 0.1) |
| 40–49 | 0.3 (0.3, 0.3) | 0.3 (0.3, 0.3) | 0.2 (0.2, 0.3) | 0.2 (0.2, 0.3) | 0.1 (0, 0.1) | 0.1 (0, 0.1) |
| 50–59 | 0.6 (0.5, 0.6) | 0.6 (0.5, 0.6) | 0.5 (0.4, 0.6) | 0.5 (0.4, 0.6) | 0.2 (0.1, 0.4) | 0.2 (0.1, 0.4) |
| 60–69 | 1.0 (ref) | 1.0 (ref) | 1.0 (ref) | 1.0 (ref) | 1.0 (ref) | 1.0 (ref) |
| 70–79 | 2 (1.8, 2.3) | 2 (1.8, 2.3) | 1.6 (1.3, 2.1) | 1.7 (1.3, 2.2) | 6.9 (5.2, 9.5) | 7.1 (5.3, 9.7) |
| 80–89 | 1.5 (1.3, 1.8) | 1.6 (1.3, 1.9) | 0.4 (0.2, 0.6) | 0.4 (0.2, 0.6) | 23.7 (17.9, 32) | 26.7 (20, 36) |
| 90+ | 0.7 (0.5, 1) | 0.8 (0.6, 1) | 0 (0, 0) | 0 (0, 0) | 45.1 (33, 62.5) | 55.8 (40.5, 77.9) |
| Sex |  |  |  |  |  |  |
| Male |  | 1.3 (1.2, 1.4) |  | 1.9 (1.6, 2.3) |  | 1.9 (1.6, 2.2) |
| Female |  | 1.0 (ref) |  | 1.0 (ref) |  | 1.0 (ref) |

The more comorbidities a person had, the higher was their risk of severe COVID-19. The effect was even stronger when the analysis was restricted to 20–69-year-olds (Table 3b). Comparing SARS-CoV-2 cases with three or more comorbidities with cases without any of the comorbidities in question, the age- and sex-adjusted ORs of severe outcomes were 6.6 for hospitalisation (95% CI 5.6, 7.7), 9.5 for ICU admission (95% CI 7.0, 12.9), and 16.1 for death (95% CI 8.1, 33.4).

**Table 3b.** Relative risk of severe COVID-19 (hospitalisation, ICU and death) in cases of SARS-Cov-2 infection by number of comorbidities and by individual comorbidities, adjusted for age and sex. Relative risks are presented in terms of odds ratios (OR) and their 95% confidence intervals (CI 95%).

|  | OR (95% CI) |  |  |  |  |  |
| --- | --- | --- | --- | --- | --- | --- |
|  | Hospitalised |  | Hospitalised, ICU |  | Fatal cases |  |
|  | Crude | Adjusted for age and sex | Crude | Adjusted for age and sex | Crude | Adjusted for age and sex |
| **Number of comorbidities, all ages<sup>1</sup>** |  |  |  |  |  |  |
| 1 | 4.3 (3.9, 4.8) | 2.4 (2.2, 2.7) | 4.7 (3.7, 6) | 2.5 (2, 3.2) | 15.3 (11.1, 21.5) | 2.6 (1.9, 3.8) |
| 2 | 8.5 (7.6, 9.5) | 3.5 (3.1, 4) | 10.9 (8.6, 13.9) | 4.5 (3.4, 5.8) | 64.5 (47.5, 89.8) | 4.4 (3.1, 6.3) |
| 3+ | 15.6 (13.8, 17.6) | 5.3 (4.6, 6.1) | 16.7 (13, 21.5) | 6.2 (4.6, 8.2) | 146.2 (107.8, 203) | 6.2 (4.4, 9) |
| **Number of comorbidities, 20–69-year-olds<sup>1</sup>** |  |  |  |  |  |  |
| 1 | 3.5 (3.1, 3.9) | 2.5 (2.2, 2.8) | 3.7 (2.8, 4.8) | 2.4 (1.8, 3.2) | 8.6 (4.6, 17.2) | 4.7 (2.5, 9.6) |
| 2 | 6.6 (5.8, 7.6) | 3.8 (3.3, 4.4) | 10.3 (7.8, 13.6) | 5.4 (4, 7.2) | 26 (13.8, 51.7) | 9.9 (5.1, 20.3) |
| 3+ | 13 (11.1, 15.2) | 6.6 (5.6, 7.7) | 21.4 (16.1, 28.4) | 9.5 (7, 12.9) | 52.9 (27.8, 106.2) | 16.1 (8.1, 33.4) |
| **Comorbidities, 20–69-year-olds<sup>2</sup>** |  |  |  |  |  |  |
| Organ or stem cell transplant | 10.7 (6.2, 18.1) | 7.2 (4, 12.5) | 17.5 (8, 34.1) | 9.8 (4.3, 19.8) | 6.9 (0.4, 31.6) | 2.8 (0.2, 13.5) |
| Malignant neoplasms | 3.4 (2.7, 4.3) | 1.7 (1.4, 2.2) | 3.1 (1.8, 4.8) | 1.4 (0.9, 2.3) | 14.8 (8.6, 24.3) | 5.8 (3.3, 9.7) |
| Hematologic malignancy | 5 (3.1, 7.8) | 2.9 (1.7, 4.6) | 6.4 (2.7, 12.9) | 3.1 (1.3, 6.4) | 15.2 (4.6, 37.1) | 5.6 (1.6, 14.1) |
| Severe disorders of the immune system | 7.4 (3.4, 14.9) | 8 (3.5, 17.2) | 2.7 (0.2, 12.6) | 2.5 (0.1, 12.4) | 0 (0, 5211.9) | 0 (0, 4720.9) |
| Severe chronic renal disease | 10.5 (7.5, 14.7) | 6.7 (4.6, 9.6) | 12.5 (7.2, 20.2) | 6.1 (3.5, 10.2) | 27.2 (12.5, 52.2) | 9.4 (4.2, 18.8) |
| Severe chronic liver disease | 3.1 (1.4, 6) | 1.5 (0.7, 3) | 4.6 (1.1, 12.5) | 2.1 (0.5, 5.7) | 26.9 (8.1, 67) | 9.8 (2.9, 25.7) |
| Asthma | 3.3 (2.9, 3.8) | 3 (2.5, 3.4) | 4.9 (3.8, 6.3) | 4.3 (3.3, 5.6) | 2.6 (1.3, 4.6) | 2 (1, 3.7) |
| Chronic pulmonary disease | 8.8 (6, 12.9) | 3.7 (2.5, 5.5) | 10.7 (5.5, 18.7) | 3.8 (1.9, 6.7) | 21.7 (8.3, 46.5) | 5.4 (2, 11.9) |
| Primary or secondary hypertension | 5.1 (4.5, 5.7) | 2.3 (2.1, 2.7) | 7 (5.7, 8.7) | 2.9 (2.3, 3.7) | 7.9 (5.1, 11.8) | 2.2 (1.4, 3.4) |
| Essential (primary) hypertension alone | 1.7 (1.3, 2.2) | 0.8 (0.6, 1.1) | 1.3 (0.6, 2.4) | 0.6 (0.3, 1.1) | 1.2 (0.2, 3.8) | 0.5 (0.1, 1.5) |
| Secondary hypertension (hypertensive heart and/or renal disease) alone | 5 (4.2, 5.9) | 2.1 (1.7, 2.5) | 8.5 (6.4, 11.2) | 3.2 (2.4, 4.3) | 10.6 (6.2, 17.1) | 2.7 (1.6, 4.5) |
| Ischaemic heart diseases | 5.2 (4.2, 6.5) | 2 (1.6, 2.6) | 7.3 (4.9, 10.3) | 2.4 (1.6, 3.4) | 9.9 (5, 17.9) | 2.1 (1, 3.9) |
| Heart failure | 4.3 (2.2, 7.7) | 2.3 (1.1, 4.3) | 3 (0.5, 9.6) | 1.3 (0.2, 4.1) | 6.1 (0.3, 28.1) | 1.8 (0.1, 8.6) |
| Type 1 diabetes | 1.9 (1.3, 2.8) | 1.9 (1.2, 2.8) | 1.6 (0.6, 3.4) | 1.4 (0.5, 3.1) | 1.3 (0.1, 5.7) | 1 (0.1, 4.8) |
| Type 2 diabetes | 5.8 (5, 6.6) | 2.8 (2.4, 3.3) | 8.5 (6.6, 10.8) | 3.7 (2.8, 4.7) | 10.1 (6.3, 15.7) | 3.2 (2, 5.1) |
| Diseases of myoneural junction and muscle | 7.7 (4.2, 13.6) | 5.5 (2.9, 10.2) | 13.8 (5.7, 28.7) | 8.1 (3.2, 17.9) | 14.9 (2.4, 48.7) | 5.8 (0.9, 20.4) |
| Cerebral palsy and other paralytic syndromes | 3.6 (1.8, 6.6) | 3.3 (1.6, 6.2) | 5.8 (1.8, 14.1) | 4.9 (1.4, 12.4) | 5.8 (0.3, 26.4) | 4.6 (0.3, 22.6) |
| Cerebrovascular diseases | 5.5 (4, 7.4) | 2.5 (1.8, 3.4) | 6.3 (3.6, 10.4) | 2.4 (1.3, 4) | 14.3 (6.3, 28) | 3.8 (1.7, 7.6) |
| Sleep apnea | 4.1 (3.6, 4.8) | 2.2 (1.9, 2.6) | 5.6 (4.3, 7.3) | 2.7 (2, 3.5) | 3.4 (1.7, 6.1) | 1.2 (0.6, 2.2) |
| Psychotic disorders | 3.4 (2.5, 4.6) | 3.4 (2.5, 4.7) | 1.3 (0.5, 2.9) | 1.2 (0.4, 2.7) | 11 (5.1, 20.8) | 10.2 (4.6, 20) |
| Clozapine (prescription for) | 4.3 (2.1, 8) | 4.2 (2, 8) | 3.3 (0.5, 10.6) | 2.9 (0.5, 9.7) | 6.7 (0.4, 31) | 6.1 (0.3, 30.6) |
| Glucocorticoids (prescription for) | 3.3 (2.9, 3.7) | 2.6 (2.3, 2.9) | 3.2 (2.5, 4.1) | 2.4 (1.8, 3) | 4.3 (2.7, 6.7) | 3 (1.9, 4.7) |
| Biologic drugs (entitlement to reimbursement of medical expenses) | 1.8 (1, 3) | 1.7 (0.9, 2.9) | 0.6 (0, 2.6) | 0.5 (0, 2.4) | 0 (0, 0.8) | 0 (0, 76.3) |
| Biologic drugs, Tumor necrosis factor alpha (TNF-α) inhibitors (prescription for) | 1.4 (0.6, 2.6) | 1.4 (0.6, 2.6) | 0.7 (0, 3.2) | 0.7 (0, 3.3) | 0 (0, 2.4) | 0 (0, 462.3) |
| Biologic drugs, other (prescription for) | 3.7 (1.6, 7.4) | 2.9 (1.2, 5.9) | 1.9 (0.1, 8.9) | 1.4 (0.1, 6.5) | 0 (0, 46.4) | 0 (0, 0) |
| Other Immunosuppressant drugs | 3.3 (2.5, 4.2) | 2.5 (1.9, 3.3) | 3.3 (1.9, 5.4) | 2.4 (1.4, 3.9) | 3.4 (1, 8.1) | 2.2 (0.7, 5.3) |
| Autoimmune disease with immunosuppressive drug therapy | 3 (2.4, 3.8) | 2.3 (1.8, 2.9) | 2.5 (1.5, 4) | 1.8 (1.1, 2.8) | 4.8 (2.1, 9.3) | 3.1 (1.4, 6.1) |
| Autoimmune disease without immunosuppressive drug therapy | 1.7 (1.2, 2.2) | 1.2 (0.9, 1.6) | 2 (1.1, 3.3) | 1.4 (0.8, 2.4) | 0.6 (0, 2.7) | 0.4 (0, 1.7) |

<sup>1</sup> Compared to the individuals without any of the comorbidities in question. <sup>2</sup> Comparing the odds of each outcome with and without the comorbidities in question.

Many of the individual comorbidities were clear risk factors for death. Among the 20–69-year-olds the strongest risk factors for death due to COVID-19 were severe chronic renal disease (OR 9.4; 95% CI 4.2, 18.8), malignant neoplasms (OR 5.8; 95% CI 3.3, 9.7), hematologic malignancy (OR 5.6; 95% CI 1.6, 14.1), chronic pulmonary disease (OR 5.4; 95% CI 2.0, 11.9), and cerebral palsy and other paralytic syndromes (OR 4.6; 95% CI 0.3, 22.6), when comparing the odds between individuals with and without the comorbidity in question (Table 3b). Likewise, cerebrovascular diseases, type 2 diabetes, autoimmune diseases treated with immunosuppressive drugs, use of glucocorticoids, secondary hypertension alone, ischaemic heart diseases and asthma increased the risk of death, with ORs between 2 and 5. Some of the comorbidities were very rare, thus, it was not possible to calculate reliable OR estimates.

Strong risk factors for COVID-19 hospitalisation among the 20–69-year-olds were severe disorders of the immune system (OR 8.0; 95% CI 3.5, 17.2), organ or stem cell transplant (OR 7.2; 95% CI 4.0, 12.5), chronic renal disease (OR 6.7; 95% CI 4.6, 9.6), diseases of myoneural junction and muscle (OR 5.5; 95% CI 2.9, 10.2) (Table 3b). Likewise, psychotic disorders, chronic pulmonary disease, paralytic syndromes, asthma, hematologic malignancy, type 2 diabetes, cerebrovascular diseases, cardiovascular diseases, autoimmune diseases treated with immunosuppressive drugs, sleep apnea, and the use of clozapine, immunosuppressants or biologic drugs increased the risk of hospitalisation, with ORs between 2 and 5. The severity of the comorbidity was associated with severe disease outcomes. The risk of hospitalisation and admission to ICU was higher for asthma and type 2 diabetes cases that had been treated in secondary health care (Figure 3).

**Figure 3.**
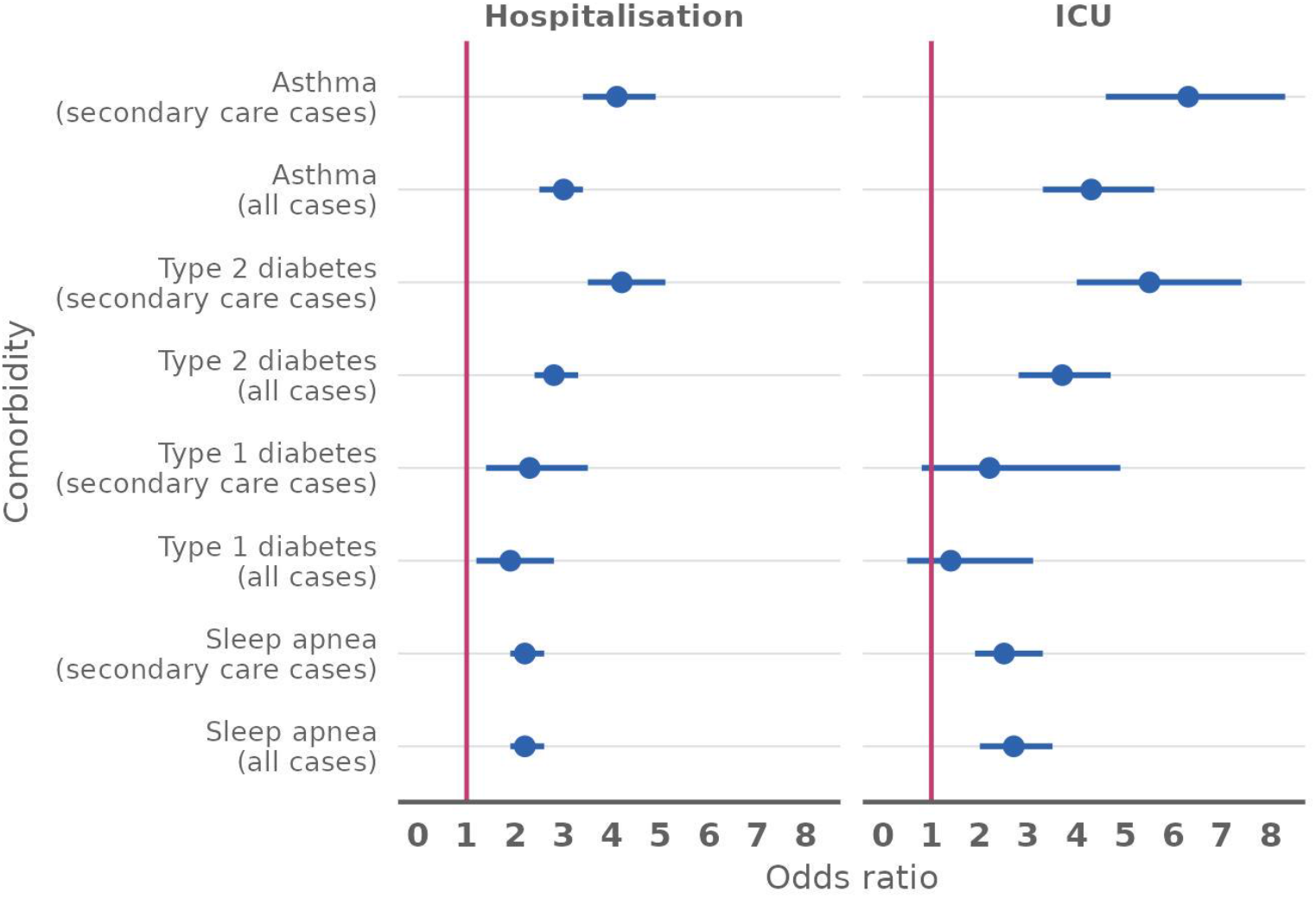
Predictors of severe COVID-19 (hospitalisation and ICU) in cases of SARS-Cov-2 infection. Odds ratios (OR) and 95% confidence intervals (CI 95%).

According to the negative binomial regression model, the population-based rate of SARS-CoV-2 infection decreased with age. Compared to the 60–69-year-olds, the rate was 3.0-times higher in 20–29-year-olds (RR 3.0; 95% CI 2.8, 3.3) and 2.4-times higher in 30–39-year-olds (RR 2.4; 95% CI 2.2, 2.5), but somewhat lower in 70–79-year-olds (RR 0.7; 95% CI 0.6, 0.7) and 80–89-year-olds (RR 0.9; 95% CI 0.9, 1). The higher rate of SARS-CoV-2 infection in the ≥90-year-olds (RR 1.5; 95% CI 1.4, 1.7) can be explained by local epidemics which occurred in nursing homes. However, the rate of SARS-CoV-2 infection did not differ between females and males. Compared to the 60–69-year-olds, the population-based rate of hospitalisation was higher among the 50–59-year-olds (RR 1.3; 95% CI 1.2, 1.4) and the risk of ICU admission was higher among the 50–59-year-olds (RR 1.2; 95% CI 1.0, 1.4).

In addition, for each comorbidity, we report the relative rates of hospitalisation, ICU admission and death in the 20–69-year-olds, comparing individuals with the comorbidity with those without any of the specified comorbidities (Table 4). The rate of SARS-CoV-2 infection was roughly the same for most of the comorbidities when comparing to those without any of the specified comorbidities (Table 4). However, some differences were obvious, e.g. those using clozapine (RR 0.5; 95% CI 0.4, 0.7), with psychotic disorders (RR 0.6; 95% CI 0.5, 0.7) and heart failure, (RR 0.7; 95% CI 0.6, 0.9) had lower rates of SARS-CoV-2 infection. By contrast, the relative rate of severe COVID-19 outcomes in the 20–69-year-olds, when comparing to those without any of the specified comorbidities varied, e.g. For hospitalisation it was 5.1–8.9-times higher for organ or stem cell transplant, severe chronic renal disease, severe disorders of the immune system, diseases of myoneural junction and muscle and type 2 diabetes.

**Table 4.** Relative rates of SARS-Cov-2 infection and severe COVID-19 outcomes (hospitalisation, admission to ICU and death) in the Finnish population aged 20–29, based on a negative binomial regression model. The table presents rate ratios (RR) and 95% confidence intervals (CI 95%).

|  | RR (95% CI) |  |  |  |
| --- | --- | --- | --- | --- |
|  | SARS-CoV-2<br>PCR-positive | Hospitalised | Hospitalised, ICU | Fatal cases |
| **Age, years** |  |  |  |  |
| 20–29 | 3.1 (2.9, 3.3) | 0.4 (0.3, 0.5) | 0.2 (0.1, 0.3) | 0.1 (0, 0.3) |
| 30–39 | 2.4 (2.3, 2.5) | 0.8 (0.7, 0.9) | 0.5 (0.4, 0.7) | 0.2 (0.1, 0.3) |
| 40–49 | 2.3 (2.2, 2.4) | 1.2 (1, 1.3) | 0.9 (0.8, 1.2) | 0.3 (0.2, 0.4) |
| 50–59 | 1.8 (1.8, 1.9) | 1.3 (1.2, 1.5) | 1.2 (1, 1.4) | 0.5 (0.4, 0.7) |
| 60–69 | 1.0 (ref) | 1.0 (ref) | 1.0 (ref) | 1.0 (ref) |
| **Sex** |  |  |  |  |
| Male | 1 (1, 1) | 1.3 (1.2, 1.4) | 1.2 (1.1, 1.3) | 2.1 (1.8, 2.3) |
| Female | 1.0 (ref) | 1.0 (ref) | 1.0 (ref) | 1.0 (ref) |
| **Comorbidities, 20–69-year-olds<sup>1</sup>** |  |  |  |  |
| Organ or stem cell transplant | 1 (0.8, 1.3) | 8.9 (5.5, 13.8) | 20.7 (9.7, 38.6) | 15.2 (0.8, 76.6) |
| Malignant neoplasms | 1 (0.9, 1.1) | 3.1 (2.3, 4) | 3.6 (2.1, 5.7) | 20.7 (10, 43.9) |
| Hematologic malignancy | 1 (0.8, 1.2) | 4.7 (2.9, 7.2) | 7.6 (3.2, 15.3) | 25.9 (7.2, 73.9) |
| Severe disorders of the immune system | 0.9 (0.6, 1.2) | 7.5 (3.7, 13.5) | 4.7 (0.3, 20.8) | NA |
| Severe chronic renal disease | 1 (0.8, 1.1) | 8.3 (6, 11.3) | 14.3 (8.2, 23.2) | 46.4 (19, 108.7) |
| Severe chronic liver disease | 0.9 (0.7, 1.1) | 2.7 (1.3, 4.9) | 4.8 (1.2, 12.7) | 38.6 (10.8, 110.3) |
| Asthma | 1.1 (1.1, 1.2) | 4.8 (4, 5.9) | 9.5 (7.1, 12.7) | 9.8 (4.3, 22.1) |
| Chronic pulmonary disease | 0.8 (0.7, 1) | 4.6 (3.1, 6.6) | 7.4 (3.9, 13) | 19.5 (6.8, 50.1) |
| Primary or secondary hypertension | 0.9 (0.9, 1) | 3.5 (2.8, 4.2) | 4.9 (3.8, 6.4) | 7.5 (4, 15) |
| Essential (primary) hypertension alone | 1 (0.9, 1.1) | 1.8 (1.3, 2.4) | 1.6 (0.8, 3) | 2.2 (0.4, 8.2) |
| Secondary hypertension (hypertensive heart and/or renal disease) alone | 0.8 (0.7, 0.9) | 2.9 (2.3, 3.7) | 5.6 (4, 7.7) | 9.1 (4.4, 19.1) |
| Ischaemic heart diseases | 1 (0.9, 1.1) | 3.4 (2.6, 4.5) | 5.2 (3.4, 7.7) | 8.9 (3.8, 20.2) |
| Heart failure | 0.7 (0.6, 0.9) | 2.8 (1.5, 4.9) | 2.4 (0.4, 7.4) | 6.6 (0.4, 33.6) |
| Type 1 diabetes | 0.8 (0.7, 0.9) | 2.5 (1.6, 3.7) | 2.6 (0.9, 5.8) | 4.1 (0.2, 20.7) |
| Type 2 diabetes | 1.1 (1.1, 1.2) | 5.1 (4.1, 6.3) | 7.9 (5.9, 10.5) | 12.8 (6.5, 26.1) |
| Diseases of myoneural junction and muscle | 0.9 (0.7, 1.1) | 7 (4, 11.3) | 17 (7.2, 34) | 31.5 (4.9, 114.8) |
| Cerebral palsy and other paralytic syndromes | 1 (0.7, 1.2) | 5 (2.5, 8.7) | 10.7 (3.3, 25.5) | 19.9 (1.1, 100.2) |
| Cerebrovascular diseases | 0.9 (0.8, 1.1) | 4.2 (3, 5.8) | 6 (3.3, 10) | 17.9 (7, 43.1) |
| Sleep apnea | 1 (0.9, 1) | 3.7 (3, 4.6) | 5.6 (4.1, 7.6) | 6.1 (2.6, 13.7) |
| Psychotic disorders | 0.6 (0.5, 0.7) | 3.1 (2.2, 4.1) | 1.7 (0.6, 3.7) | 24.6 (10.2, 57.2) |
| Clozapine (prescription for) | 0.5 (0.4, 0.7) | 2.9 (1.5, 5) | 3 (0.5, 9.5) | 12.7 (0.7, 63.7) |
| Glucocorticoids (prescription for) | 1.1 (1, 1.1) | 3.9 (3.2, 4.7) | 5.1 (3.8, 6.8) | 11.5 (6, 23.2) |
| Biologic drugs (entitlement to reimbursement of medical expenses) | 0.9 (0.8, 1.1) | 2.4 (1.3, 4) | 1 (0.1, 4.6) | NA |
| Biologic drugs, Tumor necrosis factor alpha (TNF-α) inhibitors (prescription for) | 0.9 (0.8, 1.1) | 1.9 (0.9, 3.5) | 1.3 (0.1, 5.7) | NA |
| Biologic drugs, other (prescription for) | 1 (0.8, 1.3) | 4.7 (2.1, 8.9) | 3.4 (0.2, 15) | NA |
| Other Immunosuppressant drugs | 0.8 (0.8, 0.9) | 3.3 (2.4, 4.3) | 4.4 (2.5, 7.3) | 7.9 (2.2, 22.3) |
| Autoimmune disease with immunosuppressive drug therapy | 0.9 (0.8, 0.9) | 3.1 (2.4, 4.1) | 3.6 (2.1, 5.7) | 11.4 (4.5, 27.2) |
| Autoimmune disease without immunosuppressive drug therapy | 0.9 (0.8, 0.9) | 1.9 (1.4, 2.6) | 2.8 (1.5, 4.8) | 1.6 (0.1, 8) |

<sup>1</sup>Comparing the rate of each outcome in individuals with the comorbidity in question to those without any of the specified comorbidities

At the upper part of Table 4, 60–69-year-olds is the reference category, and the rate ratios presented for each comorbidity and outcome at the bottom part of the table can be taken to present relative rates in that age group. The additional risk caused by a specific comorbidity at any other age category can be obtained by multiplying the rate ratio of interest at a lower part of the table by the rate ratio of the age group of interest. For almost all comorbidities, the relative risks of dying for 50–59-year-olds with comorbidities, were greater than for 60–69-year-olds without any comorbidities. For example, the risk for 55-year-old person with chronic pulmonary disease to die of COVID-19 is almost 10 times (0.5 × 19.5) of that of 65-year-old person without any of the listed conditions.

## Discussion

In this population-based setting we estimated the risks for four entities: SARS-CoV-2 infection and COVID-19 related hospitalisation, ICU admission and death by age and various comorbidities or conditions. Age and certain comorbidities were found to be predictors of severe COVID-19 in cases of SARS-CoV2 infection. Old age was strongly associated with mortality, and having multiple comorbidities was associated with both mortality and hospitalisation. The estimation of population-based rates of hospitalisation and death related to SARS-CoV-2 infection allowed an even more detailed characterisation of the relative roles of age and comorbidities as risk factors for severe COVID-19. Based on this analysis, vaccination of the elderly aged ≥70 years was prioritised in Finland, followed by vaccination of those with highly predisposing medical conditions.[16]

As expected, immunosuppressive states such as organ or stem cell transplantation, severe disorders of the immune system or severe renal disease were associated with increased risks of hospitalisation and ICU admission due to COVID-19. Similar results were obtained in a population-based Danish study where organ transplantation and severe kidney disease were the two medical conditions associated with the highest risks of severe COVID-19.[7] In an English study, which recorded nearly 11 000 deaths in a cohort of more than 17 million SARS-CoV-2 positive patients, organ transplantation was associated with the highest risk of death due to COVID-19 (hazard ratio 6.0; 95% CI 4.7, 7.6) among the studied medical conditions, after the analysis was adjusted for age and sex.[11] Furthermore, in our study patients with an autoimmune disease, those receiving immunosuppressive drug therapy were more prone to severe outcomes than patients without such therapy. Potential reasons for the increased risk are the immunosuppressive medication itself and the increased severity of the underlying disease that necessitated an immunosuppressive drug therapy.

Among SARS-CoV-2 cases aged 20-69, the relative risk of death was higher in patients with chronic renal or liver disease, diseases of myoneural junction and muscle, cerebral palsy or other paralytic syndromes as well as in those with psychotic disorders, especially with prescription for the psychiatric medication clozapine. The association of psychiatric disorders and high COVID-19 mortality has been explained only partly by the presence of other comorbidities. [17] Other reasons such as socioeconomic factors, delays in treatment seeking and certain deficits in cellular immunity in schizophrenia have been implicated.[17] Although it is known that clozapine causes neutropenia and predisposes for pneumonia[18] its role in COVID-19 infections has not been assed broadly.

In our analysis, asthma, similarly to other chronic pulmonary diseases, was associated with higher risk of hospitalisation and intensive care treatment, but it was not associated with higher mortality. The association with hospitalisation was weaker but present even when asthma had been treated in primary health care only, indicating a presumably less severe manifestation. However, in a review and meta-analysis where 76 studies and more than 17 million SARS-CoV-2 positive patients were included, asthma was not associated with severe COVID-19.[19] In our study, 4.7% of 20–69-year-old SARS-CoV-2 cases were recognized as having asthma although the overall prevalence in Finland is twice as high.[16] Our estimate is likely to represent the risk for more severe asthma. In line with that, recent treatment of asthma with corticosteroids, indicating a more severe manifestation, has been reported to be associated with an increased risk of severe COVID-19 in some studies.[7,11]

Although they are no typical predictors for other infectious diseases, it has been widely noted that metabolic syndrome and obesity as well as type 2 diabetes are strongly associated with higher risks of severe COVID-19.[11,12,14,20] In our study, type 2 diabetes, which often occurs together with obesity and metabolic syndrome, was associated with higher risks of severe COVID-19 and death than the more serious disease, type 1 diabetes. In addition to obesity, regulation of blood glucose levels might play a role in the progression of SARS-CoV-2 infections among type 2 diabetes patients as high blood sugar levels have been shown to predict severe COVID-19.[11,20,21] Moreover, type 2 diabetes has been identified as an prognostic factor for survival among COVID-19 cases requiring critical care especially among the middle-aged and younger.[14]

In many studies, hypertension has been more prevalent among severe or fatal COVID-19 cases than among milder cases.[22] Also in our study, hypertension was more prevalent among severe cases; while only 7% of the SARS-CoV-2 positive 20-69-year-old individuals were hypertensive, 25%, 32% and 36% of the hospitalised, ICU and fatal 20-69-year-old cases suffered from hypertension. However, patients who had only hypertension were not at increased risk of severe outcomes according to our study; similar results have been obtained elsewhere.[23] Since hypertension is common in patients who also have other comorbidities predisposing for severe COVID-19, such as obesity, hypertension alone is not an appropriate indicator for the prioritisation of COVID-19 vaccination.

The main strength of our study is the utilisation of multiple national registers, across which individual-level data were linked via a unique personal identifier. Information on possible predisposing comorbidities could therefore be verified from different sources. Furthermore, not only the conditional risk of hospitalisation given SARS-CoV-2 infection but also the rate of SARS-CoV2 infection could be evaluated based on our nationwide dataset. It is a strength of our study that the effect of comorbidities on the risk on severe COVID-19 was analysed among the 20-69-year-olds, excluding the elderly, which should be prioritised by their age in any case.

Our study is solely based on register data. Therefore, we could not address the severity of the underlying comorbidities to any further extent than what could be deduced from the recorded diagnoses and medications. Another weakness is the rather limited number of cases, although we included all laboratory-confirmed SARS-CoV-2 cases that had occurred by 24 February 2021. During the study period, non-pharmaceutical interventions reduced the spread of COVID-19 in Finland considerably. Additionally, not all national registers are adequate for real-time use. For example, the Finnish Cancer Register is updated only annually, so that, at the time of the study, data from 2019 and beyond were not yet available. Therefore, we could not utilise more detailed data from the Cancer register on the status or treatment of cancers and, therefore, could not identify cases receiving immunosuppressive treatments. Furthermore, the national registers used to identify individuals with the specified comorbidities did not contain private or occupational health care visits. However, if any disease entitling a special reimbursement is diagnosed at those visits, it is registered in the SII and thus included in our dataset.

When vaccinations are targeted to prevent severe COVID-19 among those with very common comorbidities, the number of vaccine doses needed is reasonably high. However, if we assume that vaccination prevents all severe cases (e. g. hospitalisations) and that the whole population, without vaccinations, would eventually acquire SARS-CoV-2 infection, the number needed to vaccinate to prevent one COVID-19 hospitalisation would remain small even for common diseases: only 4.8 (1293 infections / 271 hospitalisations; Table 2) for type 2 diabetes, 6.1 for sleep apnea and 7.3 for asthma.

According to our study, vaccination of the more critically ill, for example, patients with heart failure or type 1 diabetes, is in fact slightly less effective in preventing severe COVID-19: the risks of hospitalisation, intensive care treatment and death, conditional on SARS-CoV-2 infection, are lower than among type 2 diabetes patients. Furthermore, in Finland patients with heart failure and type 1 diabetes were less likely to have a SARS-CoV-2 infection than patients with type 2 diabetes or asthma. These findings may follow from effective shielding of the most vulnerable, i.e., strict adherence to the nonpharmaceutical interventions recommended. If in the future we are in a similar situation again (new pathogen, vaccine scarcity), this kind of study is the key.

## Data Availability

By Finnish law, the authors are not permitted to share individual-level register data. The computing code is available upon request.

## Authors’ contributions

HS, TL (Tuija), TL (Toni) conceptualized the study. TL (Toni) and UB conducted the statistical analysis. TL (Tuija) reviewed the literature. HS, TL (Tuija) and KA drafted the manuscript. All authors discussed the results and contributed to the final manuscript.

